# Association of Mental Health Treatment on Outcomes in Patients with Heart Failure and Ischemic Heart Disease

**DOI:** 10.1101/2023.05.23.23290426

**Authors:** Cheryl N. Carmin, Raymond L. Ownby, Cynthia Fontanella, Danielle Steelesmith, Philip F. Binkley

**Affiliations:** The Ohio State University Wexner Medical Center, Department of Psychiatry and Behavioral Health; Nova Southeastern University, Department of Psychiatry and Behavioral Medicine; The Ohio State University Wexner Medical Center, Division of Cardiovascular Medicine

**Keywords:** anxiety, depression, heart failure, ischemic heart disease

## Abstract

**Background:** Previous studies have not found an impact of mental health treatment on outcomes in patients with heat disease. The aim of this study was to examine whether individuals who received mental health treatment for anxiety or depression after being hospitalized for ischemic disorders or heart failure had a reduced frequency of re-hospitalizations, emergency department (ED) visits or mortality compared to those who did not receive treatment.

**Methods:** A population based, retrospective, cohort design was used to examine the association between psychotherapy or antidepressant medication prescription and health service utilization and mortality in patients with coronary artery disease and comorbid anxiety and/or depression. Those receiving versus not receiving mental health treatment were compared based on the frequency of re-hospitalization, emergency department visits, and mortality.

**Results:** The study sample included 1,563 subjects who had a mean age of 50.1 years. Individuals who received both forms of mental health treatment were 87% less likely to be re-hospitalized, 73% less likely to have an emergency department visit, and 65% less likely to die from any cause.

**Conclusions:** Mental health treatment has a significant impact on outcomes in patients with cardiovascular disease consisting of reduced hospitalizations, emergency department visits and in some conditions improved survival.

---

Heart disease is a leading cause of death and an important cause of increasing healthcare costs in industrialized nations.^1^ Among the most prevalent forms of heart disease, projections suggest that heart failure will increase 48% by 2030 resulting in over 8 million adults with this condition, with costs reaching $30.7 billion. By 2030, heart failure related costs are estimated to increase by 127%.^2^ Multiple studies have demonstrated that depression is common in those with heart failure and has a significant impact on its morbidity and mortality.^3–8^ Similarly, depression has a major impact on outcomes in patients with coronary artery disease. Following Frasure-Smith and colleagues’^9^ landmark study on depression and cardiovascular disease, multiple studies have addressed the relationship between depression and coronary artery disease, and more recently anxiety has similarly been studied.^10–12^ Research regarding the relation between anxiety, coronary artery disease and heart failure is not as extensive as that on depression, although one early study showed that phobic anxiety was a risk factor.^13^ A meta-analysis found that generalized anxiety disorder independently predicted coronary artery disease while controlling for depression.^14^ Another study found that anxiety was associated with a 41% increased risk of both coronary artery disease incidence and mortality and a 35% increased risk for heart failure.^15^ More research on anxiety, coronary artery disease and heart failure may be useful in that anxiety and depression may share common etiologies.^16–18^

In addition to morbidity and mortality, cardiovascular disease with concomitant anxiety incurs significant health care costs through hospitalizations and emergency room visits. ^19^ Much like depression, anxiety is a predictor of ambulatory health care use and rehospitalization.^20,21^ A recent study of patients with heart failure noted that anxiety, depression or their combination significantly contributed to all-cause mortality but did not predict rehospitalization^3^. In contrast, Suzuki and colleagues^4^ found that comorbid anxiety and depression were predictive of both re-hospitalization and mortality in patients with heart failure.

Although patients with cardiovascular disease frequently experience anxiety and depression, a Cochrane systematic review found little evidence that psychologic treatments are associated with improvement in clinical outcomes in patients with coronary artery disease or heart failure. ^22^ This was similarly true for more recent trials including ^23^ SADHEART, which found that sertraline is safe for use with patients with cardiovascular disease patients. However, this study was not designed to investigate cardiovascular disease outcomes.^24^

The aim of this study was to use a public data base to examine the impact of psychotherapy or psychopharmacologic treatments on adverse outcomes in patients admitted to the hospital with coronary artery disease or heart failure and who were diagnosed with anxiety or depression. The findings provide novel insight into the impact of mental health interventions on outcomes in patients with heart failure and coronary artery disease.

## Methods

### Design and Study Cohort

The study was approved by the Ohio State University Institutional Review Board. A retrospective cohort design examined the association between mental health treatment, health service utilization and mortality in patients with coronary artery disease or heart failure and comorbid anxiety or depression. We included all adults aged of 21 to 64 years over a three-year period who had a first hospital admission for two claims for ischemic heart disease or heart failure diagnoses (International classification of Diseases 9th Edition [ICD-9 CM] codes 410.X to 414.X, 428.X), and who had two or more claims for any anxiety disorder (i.e., ICD 9-CM 300.00, 300.1, 300.02, 300.21, 300.22, 300.23, 300.3X, 308.3X, 309.0X, 309.24, 309.28, 309.81) or depression (i.e., ICD 9 codes 296.2X, 296.3X, 304.4X, 311). All patients were continuously enrolled in Ohio’s Medicaid program during the six-month period prior to the index (first) admission. The index admission was defined as a hospital admission with: 1) a diagnosis of coronary artery disease or heart failure at the index admission and 2) no history of coronary artery disease events or heart failure in the six-month period prior to index admission. Patients diagnosed with schizophrenia or psychosis (ICD-9 295.X), bipolar disorder (ICD 9 codes 296.00-296.1, 296.4-296.8), dementia (ICD 9 codes 290-290.9), autism spectrum disorder (ICD 9 codes 299.00, 299.8, 299.9) and intellectual disability (ICD 9 codes 317-319) were excluded.

### Data Sources

Data came from two sources: Ohio Medicaid claims files and death certificate files over a three-year period. Medicaid claims data included information on outpatient, inpatient, and pharmacy service claims, demographic and clinical characteristics including up to nine ICD-9-CM diagnoses, and program eligibility. Death certificate records included social security number, last name, first name, date of birth, race/ethnicity, county of residence, and ICD-10 codes of all causes of death. Medicaid service files and death certificate data were linked by year with a deterministic, multistep algorithm based on decedent identifier, including social security number (SSN), first and last names (truncated to the first six letters), and date of birth.

### Outcome Measures

Four time to event variables were included: (1) hospital readmission; (2) emergency department (ED) visits for coronary artery disease and heart failure (ICD 9 codes 402.X-405.X, 410.X-417.X, 420.X-429.X), (3) all-cause mortality; and (4) heart disease mortality.

### Primary explanatory variables

The primary explanatory variables were any psychotherapy (ICD-9 CM 290-319), any treatment with antidepressant medication (based on pharmacy records) or their combination following the index admission. Psychotherapy was defined by the procedure codes for diagnostic interviewing (90801, 90791, 96150, 96151) and individual, group, or family psychotherapy (90804, 90806, 90808, 90810, 90812, 90814, 90853, 90857, 90847, 90849, 90839. 90840, 96152, 96153, 96154, and local HCPC codes).

### Covariates

Demographic characteristics included patient’s age at admission, race and ethnicity (non-Hispanic White, non-Hispanic Black, Hispanic, and other), gender, and Medicaid eligibility status (disabled versus poverty). Clinical characteristics were based on claims during the 6 months prior to the index hospitalization and included the presence of at least one claim for anxiety or depressive disorder (ICD 9 CM codes 291-292).

Pharmacy claims during the study period were used to classify psychotropic medications (e.g., antidepressants, mood stabilizers, antipsychotics, and anxiolytics) and cardiac medications. For analyses, cardiac medications were categorized as β-Blockers, calcium channel blockers, angiotensin-converting enzyme (ACE) inhibitors, angiotensin receptor blockers (ARBs), statins, diuretics, digoxin, alpha agonists, alpha blockers, other lipid medications other than statins, antiarrhythmics, and long-acting nitrates.

### Statistical Analyses

Patient demographics and clinical characteristics are presented using frequencies and percentages for categorical variables and mean and standard deviation for age. Additionally, these same characteristics are described by receipt of any mental health treatment during the follow up period for each of the outcome: all-cause mortality, CHD mortality, first ED visit, and re-hospitalization. Cox proportional hazards regression was used to assess the hazard for each outcome for those who received either psychotherapy, medication, their combination or no mental health treatment during the period between index admission and outcome.

Covariates used in models were selected from a larger number of possible comorbidities in an iterative process in which correlations between comorbidities and outcomes were first examined. Those with correlations associated with a probability of 0.10 or lower were included in models and then hazards were calculated. Variables with the smallest effect on outcomes were then eliminated and the model was again run. The process of eliminating nonsignificant variables and then again calculating the model was repeated several times to arrive at a final model for all-cause mortality. For consistency across outcomes, this model was used for other outcomes.

Final models include variables that in this process were significantly associated with outcomes or considered important to include on conceptual grounds (e.g., race). Three models were developed for each outcome: Model 1 included only patient demographics and disability status as well as the interaction of age with disability; Model 2 included the variables in Model 1 and included comorbid disorders; and Model 3 added all medications to Model 2. All analyses were conducted using Stata 15.1 (StataCorp, College Station, TX).

## Results

Sample characteristics are summarized in Table 1. Table 2 presents models for each outcome and results are further presented in Figure 1. For each of the outcomes except coronary artery disease related mortality, those who received some form of mental health treatment were significantly less likely to experience the outcome than those who received no mental health treatment (e.g., those individuals who received both therapy and medication were 75% less likely to be readmitted, 74% less likely to have an ED visit, and 66% less likely to die from any cause) although for all-cause mortality the effect for antidepressant alone only approached significance (*P* = .09). Effects for coronary artery disease mortality showed a similar pattern, with reduction of hazards associated mental health treatments and still further mortality reduction associated with combined treatment, although these results were not statistically significant.

**Table 1.**
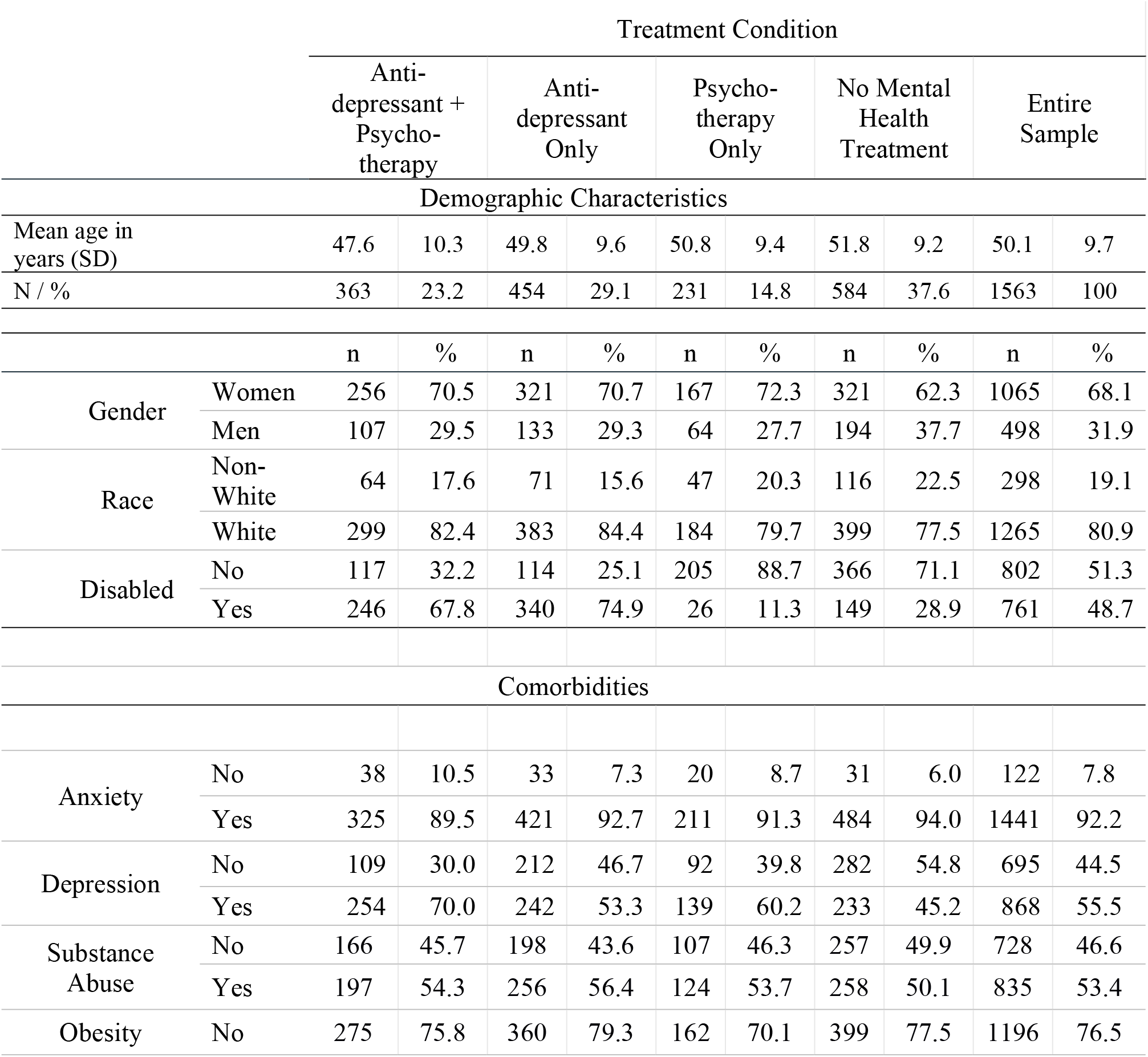
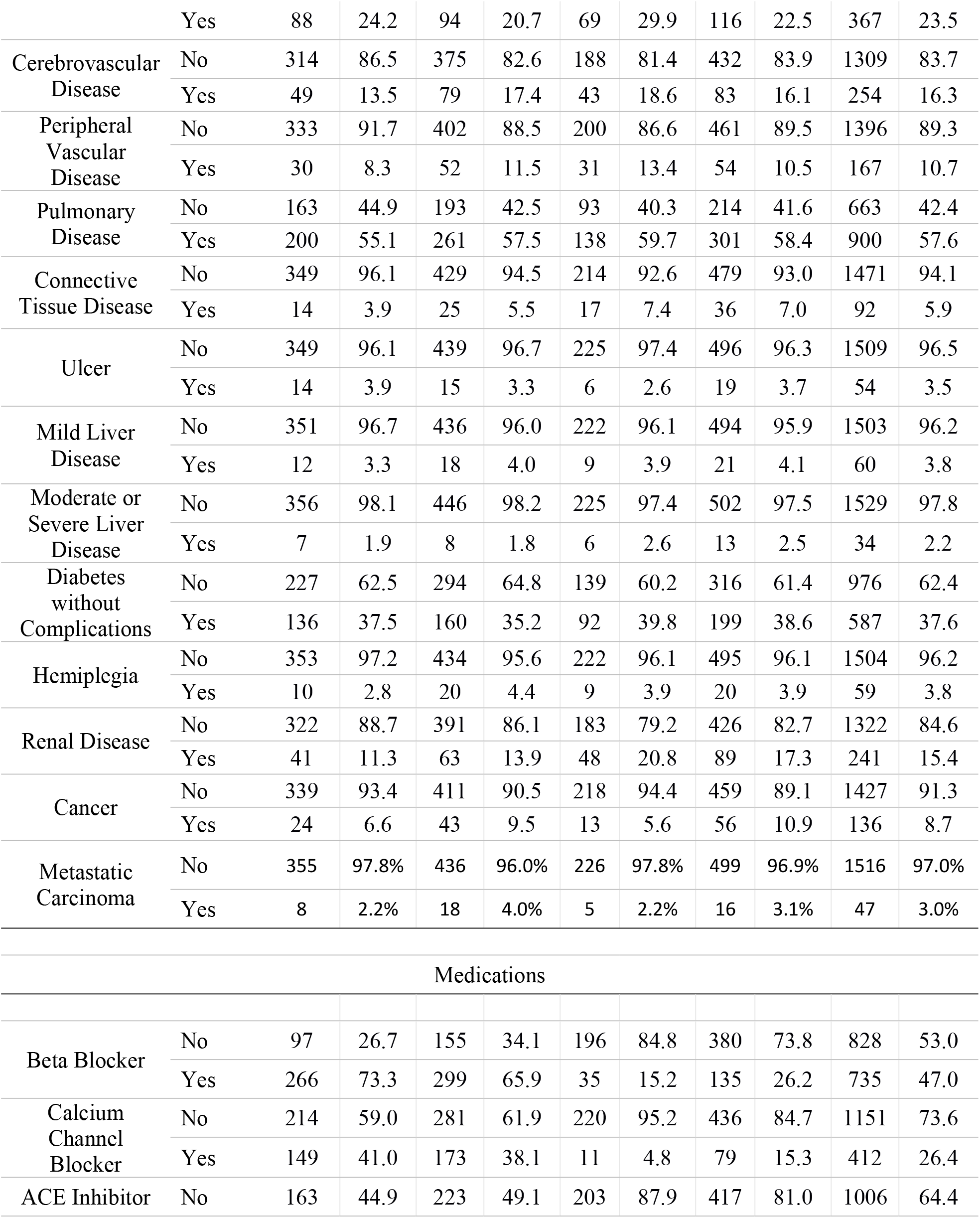
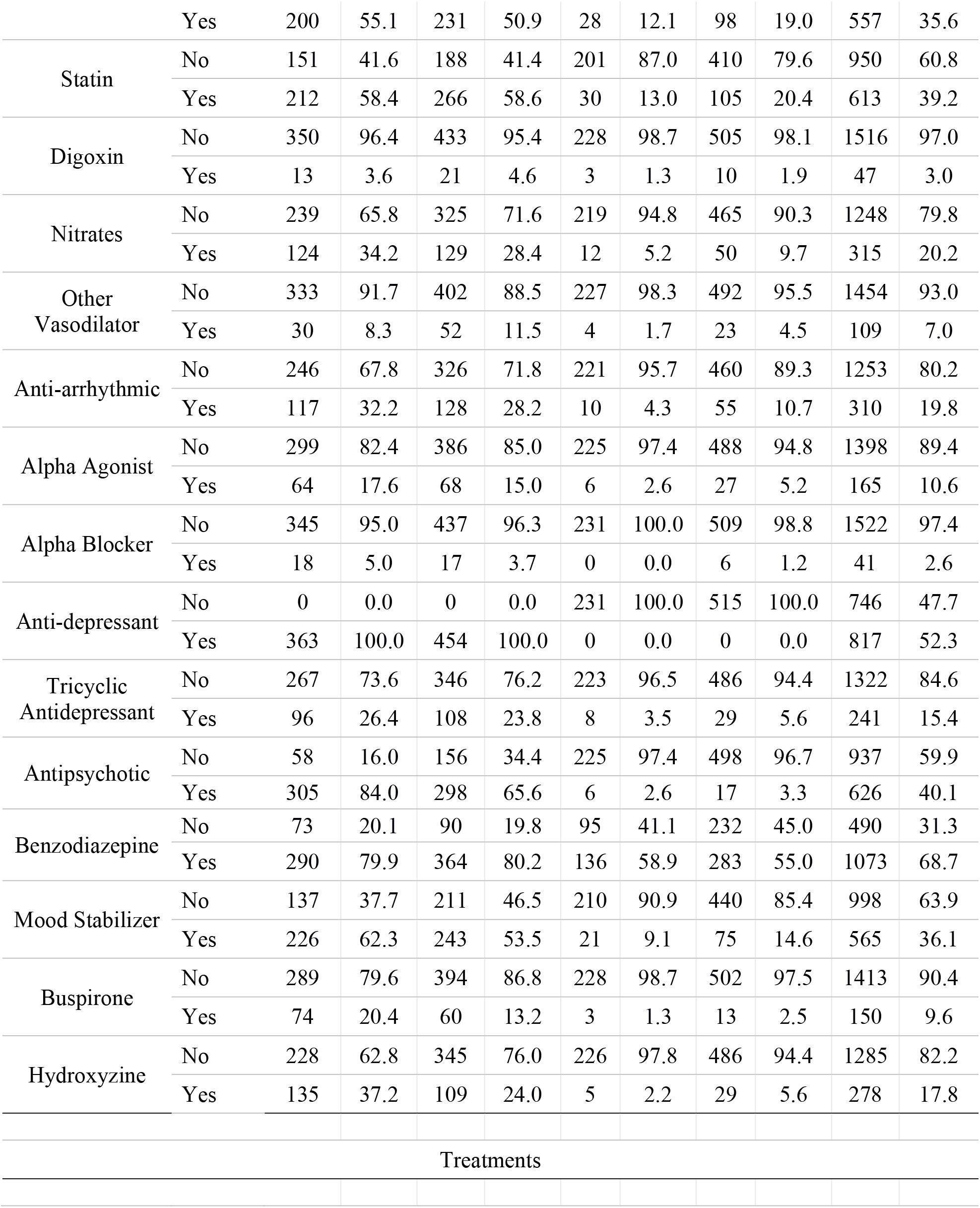
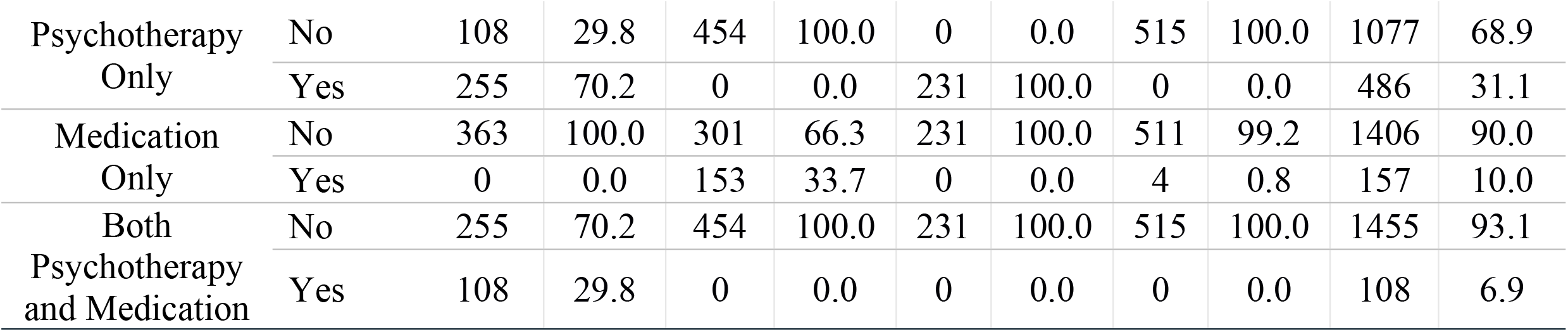
Demographics.

**Table 2.** Hazard Ratios for Cox Regression Models (N = 1,563)

|  | **Model 1<sup>a</sup>** | **Model 2<sup>b</sup>** | **Model 3<sup>c</sup>** |
| --- | --- | --- | --- |
|  | HR (95% CI), *P* | HR (95% CI), *P* | HR (95% CI), *P* |
| **All-Cause Mortality** |  |  |  |
| No treatment | 1.00 |  |  |
| Psychotherapy only | .61 (.43 - .87), .01 | .58 (.41 - .81), .002 | .52 (.37 - .75), < .001 |
| Antidepressant only | .70 (.52 - .92), .01 | .72 (.54 - .96), .03 | .70 (.47 - 1.04), .09 |
| Psychotherapy and antidepressant | .33 (.23 - .46), < .001 | .35 (.25 - .50), < .001 | .34 (.21 - .55), < .001 |
| Antidepressant only vs. psychotherapy only, *P* = > .99; Antidepressant + psychotherapy vs. antidepressant only, *P* < .001; Psychotherapy + antidepressant vs. psychotherapy only, *P* = .08; No Treatment vs. antidepressant only, *P* = .03, No treatment vs. psychotherapy only, *P* = .002; No Treatment vs. antidepressant + psychotherapy, *P* < .001.<sup>d</sup> |  |  |  |
| **Coronary Heart Disease Mortality** |  |  |  |
| No treatment | 1.00 |  |  |
| Psychotherapy only | .48 (.20 - 1.16), .10 | .47 (.21 - 1.04), .06 | .42 (.17 - 1.04), .06 |
| Antidepressant only | .92 (.48 - 1.74), .48 | .94 (.49 - 1.81), .84 | .69 (.28 - 1.72), .43 |
| Psychotherapy and antidepressant | .47 (.21 - 1.02), .06 | .47 (.19 - 1.12), .09 | .32 (.17 - 1.04), .06 |
| Antidepressant only vs. psychotherapy only, *P* = > .99; Antidepressant + psychotherapy + vs. antidepressant only, *P* = .14; Psychotherapy + antidepressant vs. psychotherapy only, *P* > .99; No treatment vs. antidepressant only, *P* => .99, No treatment vs. psychotherapy only, *P* = .18; No Treatment vs. antidepressant + psychotherapy, *P* = .11. |  |  |  |
| **Emergency Department Visits** |  |  |  |
| No treatment | 1.00 |  |  |
| Psychotherapy only | .52 (.42 - .65), < .001 | .51 (.41 - .64), < .001 | .47 (.37 - .59), < .001 |
| Antidepressant only | .58 (.50 - .67), < .001 | .59 (.50 - .68), < .001 | .51 (.42 - .62), < .001 |
| Psychotherapy and antidepressant | .32 (.26 - .39), < .001 | .33 (.26 - .40), < .001 | .26 (.20 - .34), < .001 |
| Antidepressant only vs. psychotherapy only, *P* = > .99; Psychotherapy + antidepressant vs. antidepressant only, *P* < .001; Psychotherapy + antidepressant vs. psychotherapy only, *P* = .001; No treatment vs. antidepressant only, *P* < .001, No treatment vs. psychotherapy only, *P* < .001; No Treatment vs. antidepressant + psychotherapy, *P* < .001. |  |  |  |
| **Hospital Readmission** |  |  |  |
| No treatment | 1.00 |  |  |
| Psychotherapy only | .54 (.43 - .67), < .001 | .52 (.42 - .65), < .001 | .51 (.41 - .64), < .001 |
| Antidepressant only | .53 (.45 - .62), < .001 | .53 (.45 - .63), < .001 | .42 (.34 - .52), < .001 |
| Psychotherapy and antidepressant | .30 (.24 - .38), < .001 | .32 (.26 - .40), < .001 | .25 (.19 - .34), < .001 |

Antidepressant only vs. psychotherapy only, $P = .56$ ; Psychotherapy + antidepressant vs. antidepressant only, $P < .001$ ; Psychotherapy + antidepressant vs. psychotherapy only, $P < .001$ ; No Treatment vs. antidepressant only, $P < .001$ , No treatment vs. psychotherapy only, $P < .001$ ; No Treatment vs. antidepressant + psychotherapy, $P < .001$ .
<sup>a</sup> Model 1: Adjusted for age, gender, race, disability, and the interaction age with disability. <sup>b</sup> Model 2: Model 1 plus comorbid disorders: pulmonary disease, mild liver disease, moderate liver disease, diabetes without complications, diabetes with complications, hemiplegia, renal disease, any cancer, and any metastatic carcinoma. <sup>c</sup> Model 3: Model 2 plus medication use and prior diagnosis of depression or substance abuse disorder. Medications include: ACE inhibitors, angiotensin receptor blockers, beta blockers, diuretics, statins, nitrates, antiarrhythmics, alpha agonists, any other cardiac medication, antipsychotics, benzodiazepines, mood stabilizers, buspirone, and hydroxyzine. This model also included prior mental health treatment.
<sup>d</sup> All pairwise comparisons used Bonferroni adjustments within outcomes.

**Figure 1.**
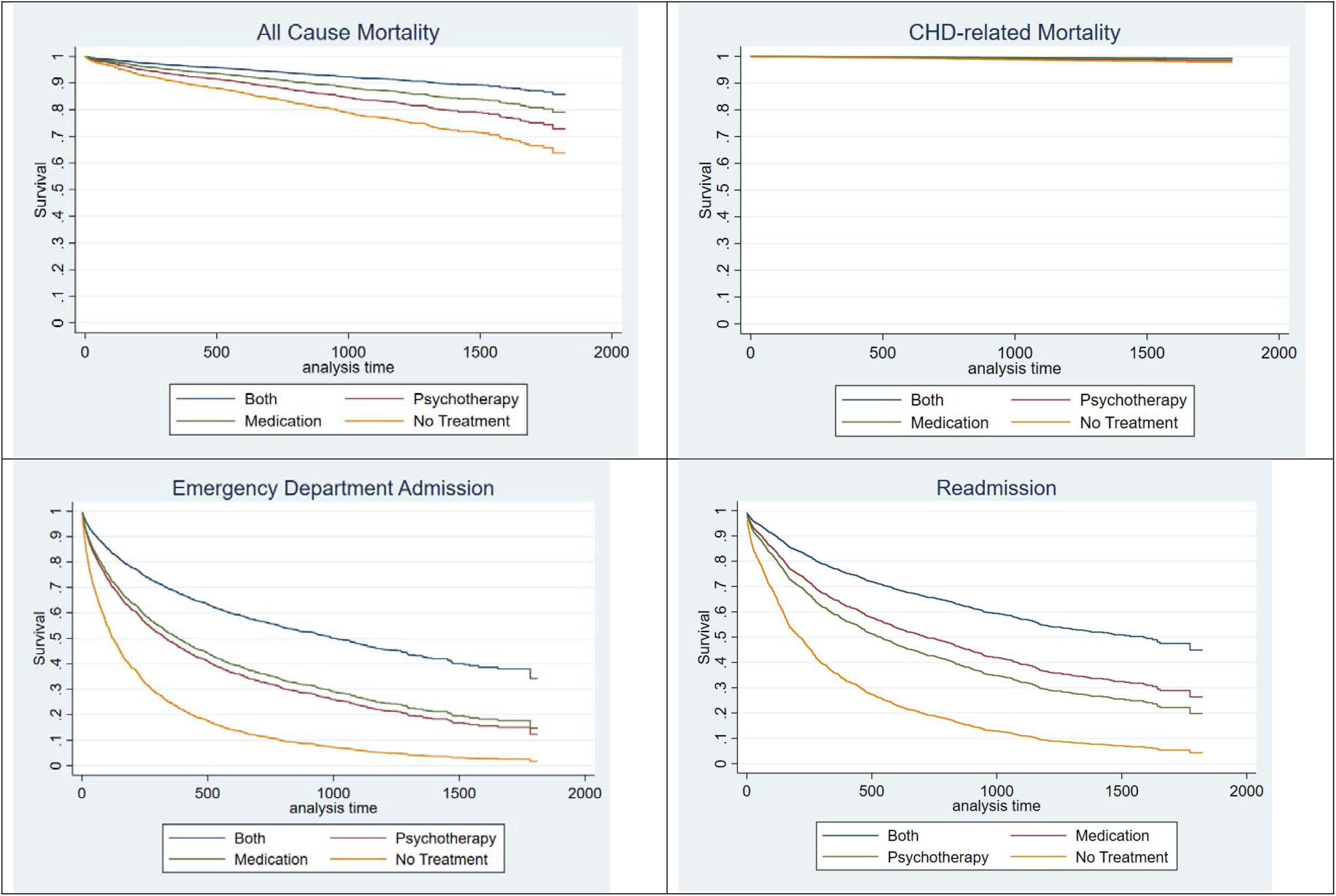
**Hazard Models** **Time to event curves for All Cause Mortality, CHD (coronary heart disease) mortality, Emergency Department admissions and Hospital Readmission with those treated with both psychotherapy and medication, only one mode of treatment and no treatment for mental health disorders. Analysis time is in days.**

## Discussion

Although there is extensive evidence that mental health has a significant impact on cardiovascular disease, there is little if any evidence that treatment of depression or anxiety has an impact on disease-related outcomes. To the authors’ knowledge, this manuscript is the first to show that mental health treatment may be associated with reduced risk for relevant outcomes. This study shows that mental health treatment, whether pharmaco- or psychotherapy, was associated with a marked and significant reduction in the risk for readmission to the hospital (75% risk reduction if receiving both treatments, 49% or 58% for psychotherapy or medication, respectively) or evaluation in the emergency room (74% if receiving both treatments, and 49% for medication treatment, and 53% for psychotherapy). These findings indicate that mental health interventions are essential to reducing hospitalizations and emergency room visits in those with heart failure or coronary disease and concomitant depression or anxiety.

Prior studies of medications for mental health treatment in patients with cardiovascular disease have largely focused on their safety. In some cases, secondary endpoints related to disease outcome have been assessed such as in SADHART-CHF^5,6^, which found a trend toward reduction in hospitalizations for patients with heart failure who received treatment for depression. Accordingly, the current study addresses a gap in the understanding of the relationship between anxiety and depression, cardiovascular disease outcomes, and treatment.

The results support the usefulness of mental health treatment in improving outcomes in patients with cardiovascular disease-related hospital admissions. They are generally consistent across models that include progressively larger numbers potential confounders, such as comorbid conditions and medications. These data suggest that treatments may be useful in reducing all-cause mortality, emergency department visits and hospital readmissions. While the findings relevant to coronary artery disease-related deaths are suggestive, they are not significant. This may be related to a relatively small sample size of patients with this diagnosis and a consequent lack of sufficient statistical power to detect an effect.

Hospital readmissions and emergency room visits for heart disease significantly contribute to the economic burden of health care in addition to diminishing patient quality of life.^25,26^ Hospitals incur Medicare monetary penalties for early readmission for some cardiovascular diseases, further contributing to costs. Interventions that can reduce the frequency of readmission and emergency room care hold the promise of significantly reducing health care costs. Considering the cost of hospital and emergency room visits versus that for mental health professional visits, our results suggest that the cost-benefit ratio for mental health care is likely to be important.

In light of recent evidence that the prevalence of anxiety exceeds that of depression^27,28^, screening patients for anxiety and depression in the clinical management of patients with heart disease is essential. Identification of patients with significant but previously unrecognized anxiety or depression allows clinicians to intervene. The data further suggest that cross disciplinary health care programs that include both cardiovascular and mental health care experts may be effective in improving outcomes and reducing costs. Effective strategies for identifying anxiety and depression in patients with subsequent effective treatment may be an important strategy by which clinicians can improve the quality of life in individuals with heart failure.^25^

The mechanisms by which mental health interventions are related to reductions in hospitalizations, emergency room visits and all-cause mortality remain speculative. However, these findings are consistent with the current understanding of the “heart brain connection.” Both heart disease and anxiety are associated with activation of the sympathetic nervous system and the production and release of proinflammatory cytokines.^27,29^ Simultaneous activation of these systems promote the progression of both CNS mediated conditions like anxiety and depression as well as heart disease. Mental health treatment may reduce both anxiety and depression and the incidence of cardiovascular events.

The study is limited in that we have sampled the Ohio Medicaid database and there is no means to ascertain if the results are generalizable to other areas of the United States or over longer periods of time. Although the specific medications used in treatment were included in the analysis, it is not possible to determine the type of psychotherapy patients received. A further limitation is that the study sample included individuals 21 to 64 years old – the age range of those covered by Medicaid. While this permits a broader sample than a Medicare cohort, it is truncated at an upper age limit that excludes older adults. Because of the observational nature of the data, we are unable to infer causality nor were we able to validate mental health diagnoses with standardized assessments. While we were able to control for a wide array of demographic and clinical confounding factors, it was also not possible to capture severity of illness or other factors that may be associated with outcomes (e.g., lifestyle factors or adherence).

In conclusion, the findings of this study demonstrate that mental health interventions for treating anxiety and depression have a significant protective impact on cardiovascular outcomes for patients with depression and anxiety. There has been question as to whether medical or person-to-person interventions, such as cognitive behavior therapy, have such an impact even though anxiety and depression are known to be important modifiers of disease risk and outcome. This study indicates that such therapies indeed have a beneficial effect on hospital and emergency department admissions and in some cases, mortality. Accordingly, these findings motivate further studies investigating mental health interventions in patients with cardiovascular disease. Studies investigating the mechanistic basis for the benefits of these interventions will guide the design of new treatment strategies that can alleviate both the personal health and financial burden of cardiovascular disease and mental illness.

### Clinical Perspective

#### What is New?

- Treatment of anxiety and depression significantly reduces hospital readmission or emergency room visits in those with heart disease who have been admitted to the hospital
- The impact of any mental health treatment on heart disease outcomes has not previously been clearly demonstrated
- The treatment of anxiety as well as depression has a significant impact on heart disease outcomes

#### What are the Clinical Implications?

- Patients with heart disease should be screened for anxiety and depression
- Those screening for anxiety and depression should undergo further diagnostic evaluation for the diagnosis of these disorders
- Patients with a definitive diagnosis of anxiety or depression should undergo appropriate treatment to improve cardiovascular outcomes as well as to treat the primary mental health diagnosis
- Collaborative care between cardiovascular experts and mental health professionals should be established to advance the care of those with heart disease

## Data Availability

All data are available to the public

## Abbreviations

ED: (emergency department)

## ACKNOWLEDGEMENTS

This study was supported by a grant from the American Psychological Association, Washington, DC to Dr. Carmin

Dr. Binkley is supported by the James W. Overstreet Chair in Cardiology at Ohio State University Wexner Medical Center.

## References

1. Davidson KW, Gidron Y, Mostofsky E, Trudeau KJ. Hospitalization cost offset of a hostility intervention for coronary heart disease patients. J Consult Clin Psychol. 2007;75(4):657–662. doi:10.1037/0022-006X.75.4.657

2. Epidemiology of heart disease and stroke - Writing group_2016.pdf.

3. Alhurani AS, Dekker RL, Abed MA, et al. The association of co-morbid symptoms of depression and anxiety with all-cause mortality and cardiac rehospitalization in patients with heart failure. Psychosomatics. 2015;56(4):371–380. doi:10.1016/j.psym.2014.05.022

4. Suzuki T, Shiga T, Kuwahara K, et al. Impact of clustered depression and anxiety on mortality and rehospitalization in patients with heart failure. J Cardiol. 2014;64(6):456–462. doi:10.1016/j.jjcc.2014.02.031

5. Jiang W, Krishnan R, Kuchibhatla M, et al. Characteristics of depression remission and its relation with cardiovascular outcome among patients with chronic heart failure (from the SADHART-CHF Study). Am J Cardiol. 2011;107(4):545–551. doi:10.1016/j.amjcard.2010.10.013

6. O’Connor CM, Jiang W, Kuchibhatla M, et al. Safety and efficacy of sertraline for depression in patients with heart failure: results of the SADHART-CHF (Sertraline Against Depression and Heart Disease in Chronic Heart Failure) trial. J Am Coll Cardiol. 2010;56(9):692–699. doi:10.1016/j.jacc.2010.03.068

7. Emani S, Binkley PF. Mind-body medicine in chronic heart failure: a translational science challenge. Circ Heart Fail. 2010;3(6):715–725. doi:10.1161/CIRCHEARTFAILURE.110.951509

8. Ferketich AK, Binkley PF. Psychological distress and cardiovascular disease: results from the 2002 National Health Interview Survey. Eur Heart J. 2005;26(18):1923–1929. doi:10.1093/eurheartj/ehi329

9. Frasure-Smith N, Lespérance F. Depression and other psychological risks following myocardial infarction. Arch Gen Psychiatry. 2003;60(6):627–636. doi:10.1001/archpsyc.60.6.627

10. Frasure-Smith N, Lespérance F. Depression and anxiety as predictors of 2-year cardiac events in patients with stable coronary artery disease. Arch Gen Psychiatry. 2008;65(1):62–71. doi:10.1001/archgenpsychiatry.2007.4

11. Frasure-Smith N, Lespérance F, Talajic M, et al. Anxiety sensitivity moderates prognostic importance of rhythm-control versus rate-control strategies in patients with atrial fibrillation and congestive heart failure: insights from the Atrial Fibrillation and Congestive Heart Failure Trial. Circ Heart Fail. 2012;5(3):322–330. doi:10.1161/CIRCHEARTFAILURE.111.964122

12. Janszky I, Ahnve S, Lundberg I, Hemmingsson T. Early-onset depression, anxiety, and risk of subsequent coronary heart disease: 37-year follow-up of 49,321 young Swedish men. J Am Coll Cardiol. 2010;56(1):31–37. doi:10.1016/j.jacc.2010.03.033

13. Kawachi I, Sparrow D, Vokonas PS, Weiss ST. Symptoms of anxiety and risk of coronary heart disease. The Normative Aging Study. Circulation. 1994;90(5):2225–2229. doi:10.1161/01.cir.90.5.2225

14. Barger SD, Sydeman SJ. Does generalized anxiety disorder predict coronary heart disease risk factors independently of major depressive disorder? J Affect Disord. 2005;88(1):87–91. doi:10.1016/j.jad.2005.05.012

15. Emdin CA, Odutayo A, Wong CX, Tran J, Hsiao AJ, Hunn BHM. Meta-Analysis of Anxiety as a Risk Factor for Cardiovascular Disease. Am J Cardiol. 2016;118(4):511–519. doi:10.1016/j.amjcard.2016.05.041

16. Chauvet-Gélinier J-C, Trojak B, Vergès-Patois B, Cottin Y, Bonin B. Review on depression and coronary heart disease. Arch Cardiovasc Dis. 2013;106(2):103–110. doi:10.1016/j.acvd.2012.12.004

17. Cohen BE, Edmondson D, Kronish IM. State of the art review: depression, stress, anxiety, and cardiovascular disease. Am J Hypertens. 2015;28(11):1295–1302. doi:10.1093/ajh/hpv047

18. Kubzansky LD, Kawachi I, Weiss ST, Sparrow D. Anxiety and coronary heart disease: a synthesis of epidemiological, psychological, and experimental evidence. Ann Behav Med. 1998;20(2):47–58. doi:10.1007/BF02884448

19. Rodwin BA, Spruill TM, Ladapo JA. Economics of psychosocial factors in patients with cardiovascular disease. Prog Cardiovasc Dis. 2013;55(6):563–573. doi:10.1016/j.pcad.2013.03.006

20. Strik JJMH, Denollet J, Lousberg R, Honig A. Comparing symptoms of depression and anxiety as predictors of cardiac events and increased health care consumption after myocardial infarction. J Am Coll Cardiol. 2003;42(10):1801–1807.

21. Baumeister H, Haschke A, Munzinger M, Hutter N, Tully PJ. Inpatient and outpatient costs in patients with coronary artery disease and mental disorders: a systematic review. Biopsychosoc Med. 2015;9(1):11. doi:10.1186/s13030-015-0039-z

22. Whalley B, Thompson DR, Taylor RS. Psychological interventions for coronary heart disease: cochrane systematic review and meta-analysis. Int J Behav Med. 2014;21(1):109–121. doi:10.1007/s12529-012-9282-x

23. Huffman JC, Mastromauro CA, Beach SR, et al. Collaborative care for depression and anxiety disorders in patients with recent cardiac events: the Management of Sadness and Anxiety in Cardiology (MOSAIC) randomized clinical trial. JAMA Intern Med. 2014;174(6):927–935. doi:10.1001/jamainternmed.2014.739

24. Glassman AH, O’Connor CM, Califf RM, et al. Sertraline treatment of major depression in patients with acute MI or unstable angina. JAMA. 2002;288(6):701–709. doi:10.1001/jama.288.6.701

25. Trudnak T, Kelley D, Zerzan J, Griffith K, Jiang HJ, Fairbrother GL. Medicaid admissions and readmissions: understanding the prevalence, payment, and most common diagnoses. Health Aff (Millwood). 2014;33(8):1337–1344. doi:10.1377/hlthaff.2013.0632

26. Hospital Guide to Reducing Medicaid Readmissions. 2014.

27. Kessler RC, Berglund P, Demler O, Jin R, Merikangas KR, Walters EE. Lifetime prevalence and age-of-onset distributions of DSM-IV disorders in the National Comorbidity Survey Replication. Arch Gen Psychiatry. 2005;62(6):593–602. doi:10.1001/archpsyc.62.6.593

28. Scott KM, de Jonge P, Alonso J, et al. Associations between DSM-IV mental disorders and subsequent heart disease onset: beyond depression. Int J Cardiol. 2013;168(6):5293–5299. doi:10.1016/j.ijcard.2013.08.012

29. Nemeroff CB, Goldschmidt-Clermont PJ. Heartache and heartbreak--the link between depression and cardiovascular disease. Nat Rev Cardiol. 2012;9(9):526–539. doi:10.1038/nrcardio.2012.91

